# Isolation of nucleated red blood cells with intact genomic DNA from cord blood by applying G&T-seq

**DOI:** 10.1101/2025.03.17.25324037

**Authors:** Noriko Ito, Tatsuya Fujii, Kosuke Taniguchi, Yuka Okazaki, Hiroko Ogata-Kawata, Haruhiko Sago, Kenichiro Hata, Kazuhiko Nakabayashi

## Abstract

**Purpose:** Fetal cells in maternal blood are a pure source of fetal genomic DNA for noninvasive prenatal genetic testing (NIPT), if successfully isolated. We assessed whether single-cell genome and transcriptome sequencing (G&T-seq), can isolate fetal nucleated red blood cells (fNRBCs) suitable for genetic testing.

**Methods:** Using umbilical cord blood as a model, we isolated 165 single NRBC candidate cells from four samples, and 12 single lymphocytes as controls from one sample. G&T-seq was used to estimate the maturation stage of each NRBC candidate from the transcriptome data and genomic integrity was assessed using genomic sequencing data.

**Results:** Multi-dimensional scaling (MDS) of the transcriptome data revealed that five NRBC candidate cells clustered separately, classifying them as primitive NRBCs. Two of these cells showed high whole-genome sequencing yields and mapping rates comparable to control lymphocytes, suggesting intact nuclear genome.

**Conclusions:** G&T-seq effectively identified primitive NRBCs with high-quality DNA among candidate cells dominated by mature RBCs. Single-cell multi-omics technology may advance the development of fNRBC-based NIPT.

## 1. INTRODUCTION

Cell-free fetal DNA was discovered in maternal blood in 1997. [1] Cell-free DNA (cfDNA)-based noninvasive prenatal genetic testing (NIPT) has become the standard method for prenatal screening for fetal chromosomal aneuploidy. cfNIPT can detect fetal chromosomal aneuploidy with a high negative predictive value. [2] However, this technology targets fragmented cfDNA primarily derived from placental cells (trophoblasts) rather than fetal cells, and the majority (80-95%) of cfDNA in the maternal blood is of maternal origin. [3] Due to insufficient positive predictive values for detecting fetal chromosomal aneuploidy, confirmatory diagnostic tests such as chorionic villus sampling (CVS) or amniocentesis are recommended when cfNIPT detects fetal chromosomal aneuploidy. [3] The accuracy of cfNIPT is also affected by several underlying maternal and fetal conditions such as autoimmune diseases, obesity, medication, malignant tumors, uterine myoma, and vanishing twins. [4-6]

Fetal nucleated red blood cells (fNRBCs) in maternal blood during pregnancy were discovered in 1959 [7] and were investigated as targets for NIPT before cfDNA. As fetal cells are the source of pure fetal genomic DNA, genetic diagnoses using genomic DNA from circulating fetal cells in the maternal blood are considered theoretically superior to those using cfDNA for detecting genomic alterations. [8] To date, four types of circulating fetal cells have been identified in the maternal blood. [9] They include fNRBCs, fetal leukocytes, fetal progenitor cells, and trophoblasts. Fetal leukocytes and progenitor cells remain in the maternal circulation for years after delivery, posing a risk of detecting fetal cells from prior pregnancies. These cell types were excluded as targets for prenatal genetic testing in cell-based NIPT development studies. [9] However, since fNRBCs and trophoblasts are rapidly lost from the maternal circulation after delivery, [9] these two cell types are being studied as targets for cell-based NIPT. Cell-based NIPT targeting circulating trophoblasts can detect aneuploidies [10], CNVs with a resolution of 1-2 Mb, [11], and disease-causing variants through single-cell-targeted [12] or whole-genome sequencing. [13] Despite their rarity in maternal blood and the difficulty in isolating them, fNRBCs are attractive targets for cell-based NIPT because they have a much lower risk of mosaic results than trophoblasts, which can be affected by confined placental mosaicism. The feasibility of cell-based NIPT targeting fNRBCs has also recently been shown to detect pathogenic variants of hereditary hearing using whole-exome and whole-genome sequencing (WGS). [14] In these studies [10-14], isolated single trophoblasts and fNRBCs were subjected to whole-genome amplification (WGA), followed by microarray or next-generation sequencing analysis. Technical improvements in the efficiency of fetal cell isolation are expected to facilitate the clinical application of cell-based NIPT. This will contribute to increasing diagnostic accuracy and may also decrease the cost and labor of the tests.

We previously reported that the combination of fluorescence-activated cell sorting (FACS) with erythrocyte-associated surface antigen markers and Y-chromosome-specific real-time PCR efficiently isolated fNRBC candidates in male infants. [15] By conducting WGA-based WGS analysis, we were able to detect fetal genomic DNA from isolated single cells. However, this method is applicable only to male infants, and the quality of the WGA-WGS data from the obtained single cells was insufficient for genomic diagnosis, likely due to the loss and/or degradation of fNRBC genomic DNA via enucleation and/or apoptosis. We hypothesized that cells in the primitive stages of NRBC maturation retain intact genomic DNA (**Fig. 1**) and screened for primitive-stage cells based on their gene expression patterns. We introduced single-cell genome and transcriptome sequencing (G&T-seq), which enabled us to obtain both transcriptome and genomic sequencing data from a single cell. [16, 17] Referring to a previously reported dataset of single-cell RNA sequencing for fetal erythroblasts at various stages of differentiation isolated from umbilical cord blood, [18] we chose marker genes for primitive and mature stages of RBC maturation. In the present study, using umbilical cord blood as a model material, we verified the effectiveness of this G&T-seq strategy for screening NRBCs with intact genomic DNA.

**Figure 1:**
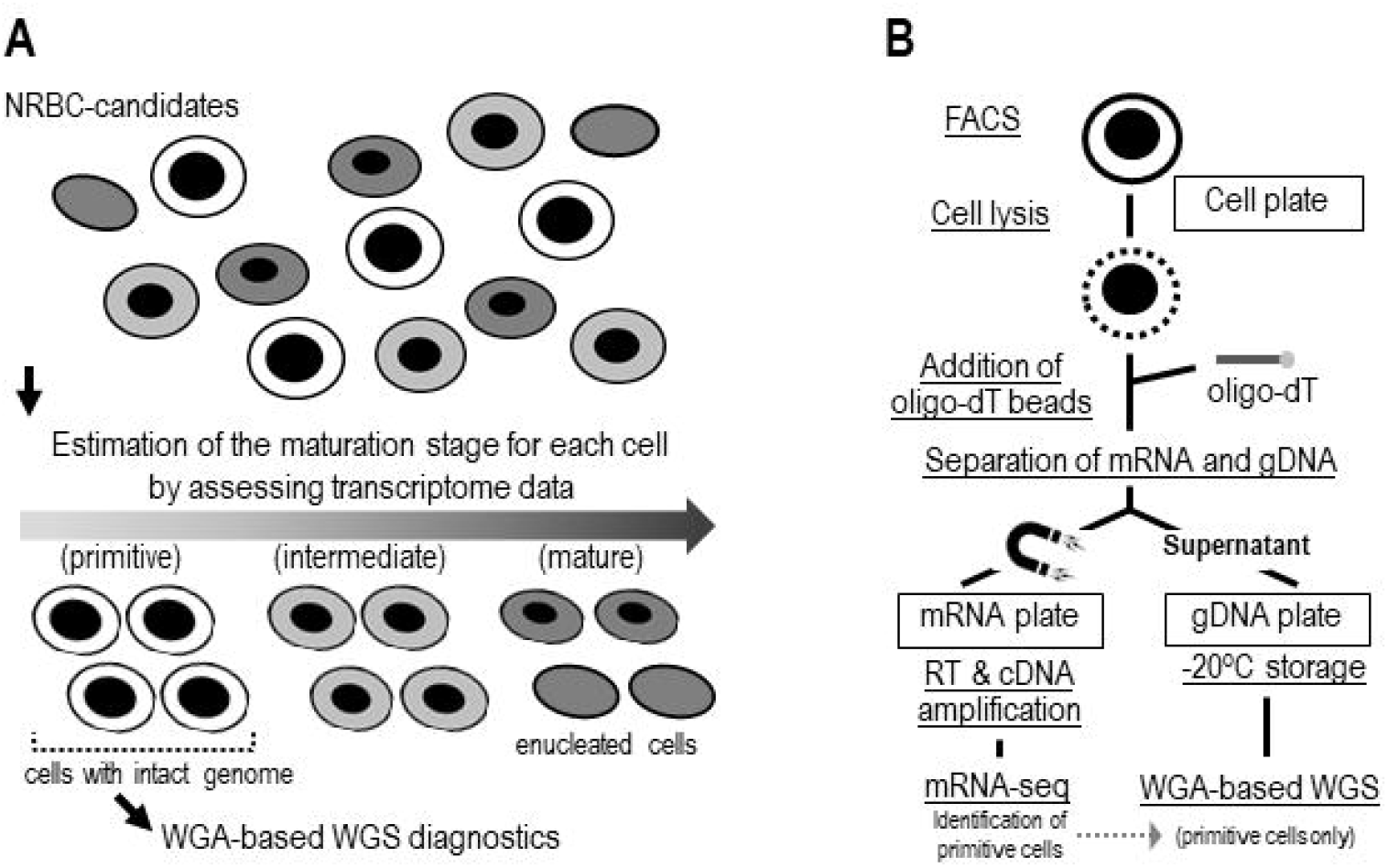
Schematic representation of the G&T-seq strategy to identify primitive-stage NRBC cells suitable for WGA-based WGS analysis. **A**. By obtaining transcriptome data for each NRBC candidate single cell and assessing the expression patterns of marker genes for primitive, intermediate, and mature stages of RBC maturation, we hypothesized that primitive-stage NRBC cells, which are expected to maintain intact genomic DNA, can be suitable for subsequent WGA-based WGS analysis. **B**. The G&T-seq workflow to first identify primitive-stage NRBCs by assessing transcriptome data and then conduct WGA-based WGS analysis only for selected cells.

## 2. MATERIALS AND METHODS

### 2.1 Blood sample collection

Adult female peripheral blood (5 mL) and umbilical cord blood (7.0 to 8.5 mL) were collected in EDTA-2K blood collection tubes.

### 2.2 immunofluorescent staining and flow cytometric analysis

Fluorescence-assisted cell sorting (FACS) was performed using a BD FACSAria III Cell Sorter (BD Biosciences). The antibodies used for FACS included BD Horizon Fixable Viability Stain 450 (BD Biosciences) to distinguish dead cells from live cells, FITC anti-human CD45 antibody (BioLegend, San Diego, CA, USA) as a leukocyte marker, and BD Pharmingen APC Mouse Anti-Human CD71 (BD Biosciences) and BD Pharmingen PE Mouse Anti-Human CD235a (BD Biosciences) as markers for erythroid precursor cells. [15] Lymphocytes from female peripheral and cord blood were sorted by gating cells positive for CD45 and negative for other markers (CD71, CCD235a, and Viability Stain 450), after gating the lymphocyte fraction in the forward scatter/side scatter plot. NRBC candidate cells were sorted from cord blood by gating cells positive for CD71 and CD235a and negative for CD45 and Viability Stain 450. Cells were sorted into the wells of a 96-well plate (#0030129512, Eppendorf, Hamburg, Germany) preloaded with 2.5 μL of Buffer RLT Plus (Qiagen, Venlo, The Netherlands). Plates containing sorted cells were stored at -80 °C.

### 2.3 G&T seq

G&T-seq was performed as previously described [17] with some modifications. In the reverse transcription step, ERCC RNA Spike-In Mix was not added, and the Maxima H Minus Reverse Transcriptase (200 U/µL) (Thermo Fisher Scientific, Waltham, MA) was used. After cDNA amplification, fragmented cDNA libraries were generated using the NEBNext Ultra II FS DNA Library Prep Kit for Illumina (New England Biolabs, Ipswich, MA). The supernatant fraction containing genomic DNA (**Fig. 2**) was purified using 0.65 volume of Agencourt AMPure XP Reagent (Beckman Coulter, Inc., Brea, CA). WGA-based WGS library preparation was conducted using the SMARTer PicoPLEX Gold Single Cell DNA-Seq Kit (Takara Bio, Shiga, Japan) according to the manufacturer’s protocol.

**Figure 2:**
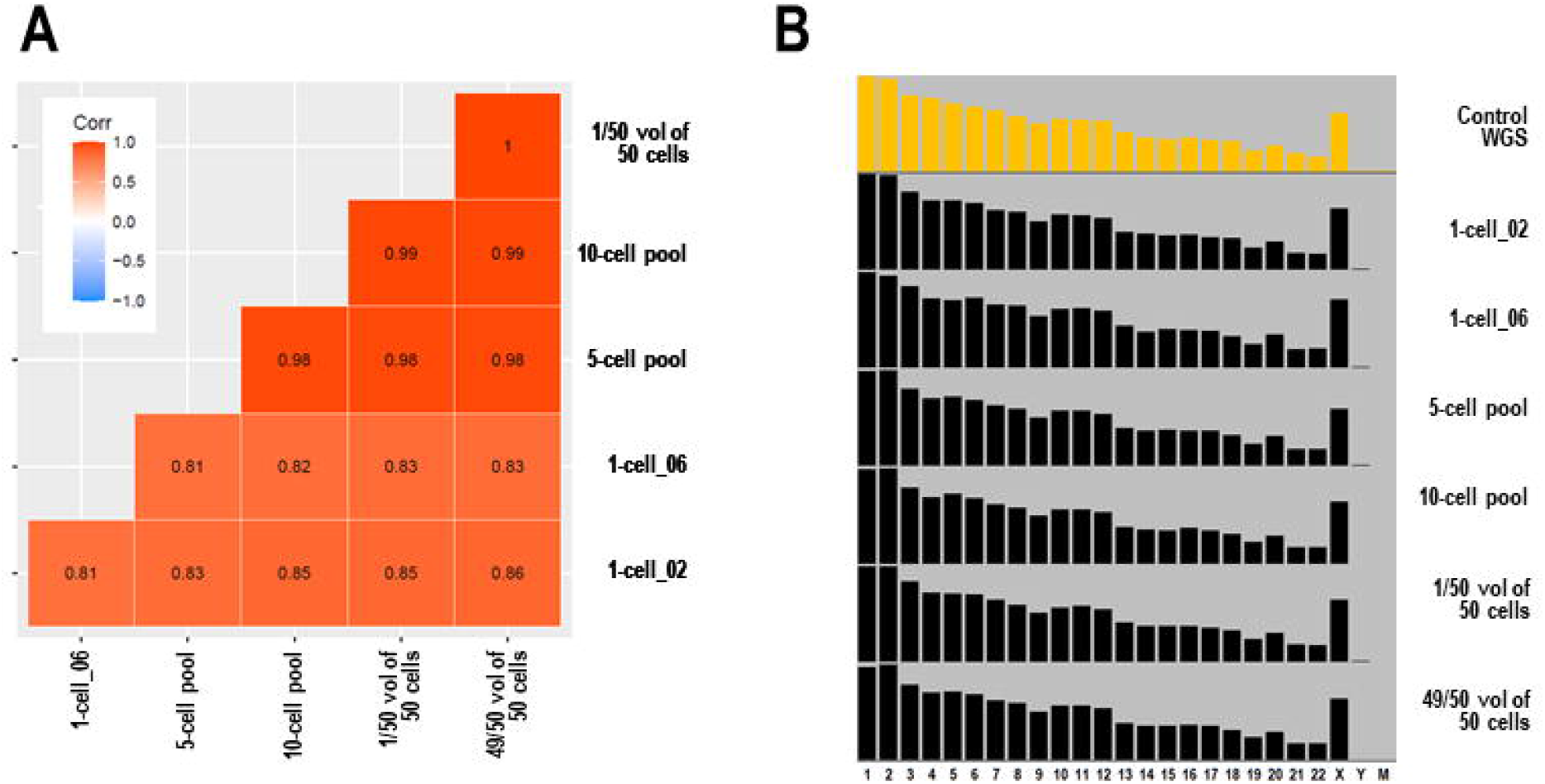
Confirmation of G&T-seq performance using single-cell lymphocytes isolated by FACS from adult female peripheral blood. RNA-seq and WGA-based WGS libraries were prepared for single cells (1-cell_02 and 1-cell_06), a five-cell pool, a 10-cell pool, a 1/50 volume of the cell lysate of 50-cell pool (“1/50 vol.of 50 cells”), and 49/50 volume of the cell lysate of 50-cell pool (“49/50 vol. of 50 cells”). **A**. The correlation coefficient matrix of transcripts per million (TPM) data of six RNA-seq libraries, whose read numbers ranging from 7.5 to 10.6 million reads (**Table S1**). **B**. Chromosomal distribution of mapped reads observed in a control WGS library generated using 600 ng of genomic DNA from female peripheral blood [15] shown in orange, and of WGA-based WGS libraries shown in blue (two one-cell libraries) and in black (other four libraries).

### 2.4 Sequencing and data analyses

RNA-seq and WGS libraries were sequenced using a MiSeq or NextSeq 550 (Illumina, San Diego, CA, USA) with paired-end (150 or 75 bp) and dual-index settings. RNA sequencing data were subjected to gene-level quantification using Salmon v1.5.2 [19] and Homo_sapiens.GRCh38v38.gtf. The correlation matrix of gene expression data was generated using the R package *ggcorrplot* with transcript-per-million (TPM) data. The Multi-dimensional scaling (MDS) plot and heatmap for the TPM values of the selected genes were generated using the R packages *EdgeR* and *gplots*, respectively. [20]

WGS data were subjected to the DRAGEN (Dynamic Read Analysis for GENomics) Bio-IT Platform v3.9.5 (Illumina, San Diego, CA) to align sequencing reads to the GRCh38 reference genome with decoy sequences. Mapping, PCR duplicate rates, and mean read depths were obtained from a report file generated by DRAGEN. The number of uniquely mapped reads for each chromosome was counted from the BAM file using the *view* command in Samtools (http://www.htslib.org). Correlation coefficients of the chromosomal distributions of the aligned genomic sequencing reads were calculated using the *correl* function in Excel.

## 3. RESULTS

### 3.1 G&T-seq for lymphocyte cells to confirm its performance

We initially tested whether we could obtain high-quality RNA-seq and WGS data by applying G&T-seq to lymphocytes isolated from adult female peripheral blood. For RNA-seq libraries, we assessed the cDNA yield and the number of genes expressed. First, the cDNA yields were assessed for the following numbers of cells (s) and wells (s): seven wells for one event (corresponding to one cell) per well, three wells for five events per well, ten events per well, and 50 events per well (16 wells in total). The cDNA yields were approximately proportional to the number of input cells. The average (and standard deviation) values were 1.2 ng (0.74 ng) for one cell, 3.5 ng (2.34 ng) for five cells, 7.8 ng (0.52 ng) for ten cells, and 22.4 ng (7.42 ng) for fifty cells (**Table S1**). We subsequently prepared RNA-seq libraries from five cDNA samples: two for one cell and one each for five cells, ten cells, and 50 cells, sequenced these libraries (8-10 million reads), and quantified the gene expression levels. The number of genes detected to be expressed (transcripts per million (TPM) > 0) was confirmed to increase proportionally with the input cell numbers: 3,962 and 6,968 for one cell, 10,125 for five cells, 12,392 for ten cells, and 16,655 for 50 cells. In the correlation analysis of the TPM data from the five RNA-seq libraries, the correlation coefficients ranged from 0.81 to 0.99 (**Fig. 2A**). Although the number of detected genes was lower in the RNA-seq libraries from one cell than in the libraries from ten and 50 cells, the single-cell libraries were expected to provide information for a sufficient number of genes to estimate the input cell types.

For WGA-based WGS libraries, we assessed library yields, PCR duplicate rates, mapping rates, and the chromosomal distribution of mapped reads. In contrast to the results of the RNA-seq libraries, the yield of the WGA-based WGS library was the highest when generated from one cell. The yields of the libraries generated from 5, 10, and 49 cells gradually decreased in proportion to the input cell number (**Table S1**). This was presumably because the WGA kit used was optimized for five cells as input. A WGS library generated from 600 ng of genomic DNA from female peripheral blood was used as a control library, showing a mapping rate of 99.2% and a PCR duplication rate of 16.3% when 836 million reads were aligned to the GRCh38 reference genome. The mapping rates of the WGA-based WGA libraries ranged from 97.85% to 99.19%. The chromosomal distribution patterns of the mapped reads of the six WGA-based WGS libraries from lymphocytes were highly concordant with those of the control WGS library (**Fig. 2B**). Correlation coefficients with those of the control WGS library ranged from 0.991 to 0.997 (**Table S1**), indicating unbiased amplification during the WGA procedure. These data demonstrate that the G&T-seq protocol we adopted enabled the satisfactory acquisition of whole transcriptome and genomic information from single cells when intact cells were subjected to library preparation.

### 3.2 G&T-seq for NRBC candidate single cells

We hypothesized that NRBCs at the primitive stages of RBC maturation contain intact genomic DNA. We conducted G&T-seq on 165 NRBC candidate cells from the umbilical cord blood (C1 to C4 samples) and 12 single lymphocytes from the C1 sample. We observed different size distributions of cDNA between lymphocytes and NRBC candidate cells. While lymphocyte cDNA showed a smear distribution as expected, NRBC candidate cells showed a sharp peak at approximately 650 bp (**Fig. 3A**). Lymphocytes and NRBC candidate cells also exhibited a remarkable difference in the top-ranked highly expressed genes: many ribosomal protein genes were top-ranked for lymphocytes, while globin-related genes were top-ranked for NRBC candidate cells (data not shown). These results demonstrated that lymphocytes and NRBCs can be distinguished by their differences in cDNA size distribution and gene expression patterns when subjected to single-cell transcriptome analysis (**Table S1**).

**Figure 3:**
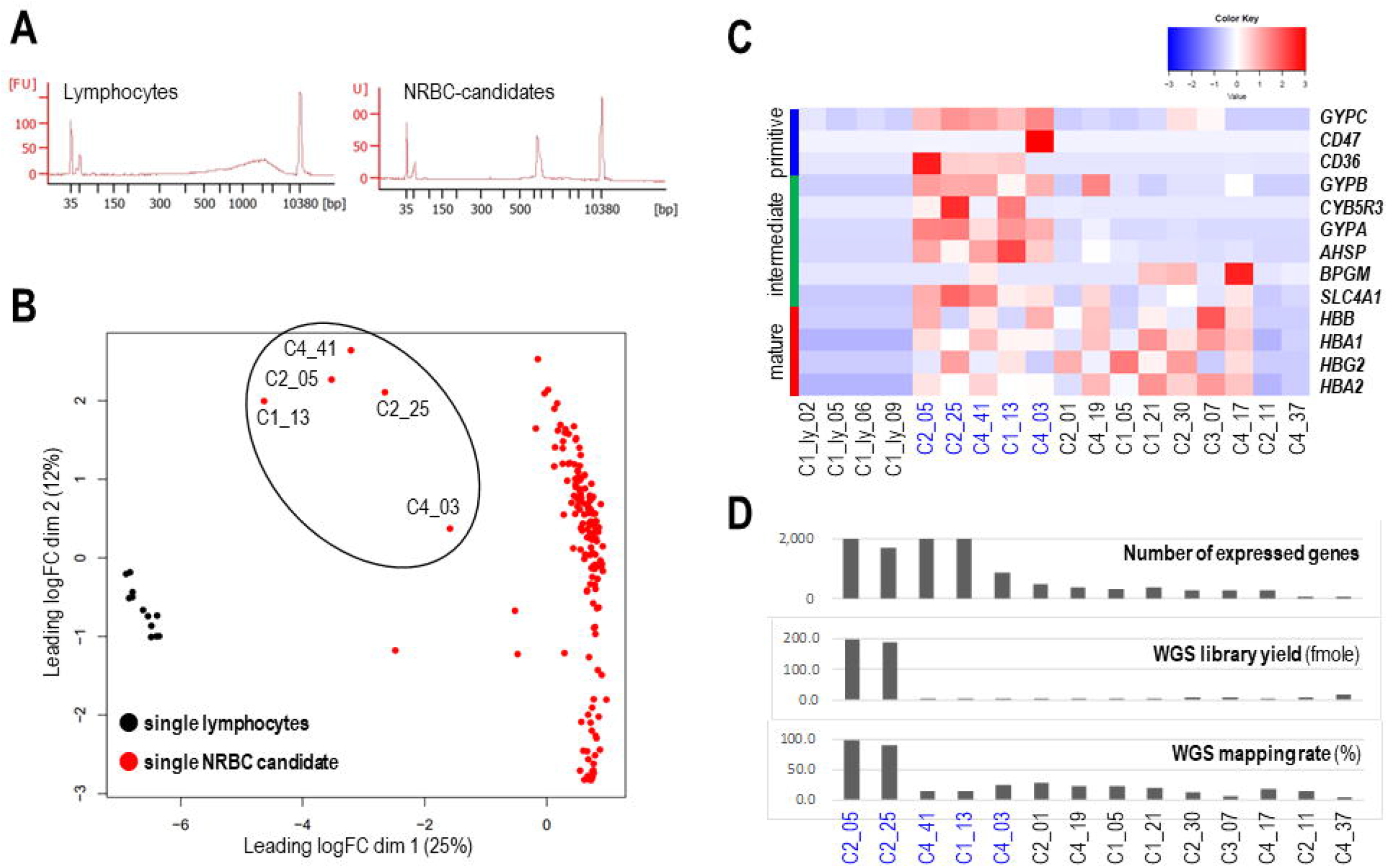
Identification of NRBCs in the primitive stage of their maturation by assessing G&T-seq data for NRBC candidates from umbilical cord blood. **A**. Size distribution of complementary DNA (cDNA) generated for RNA-seq library preparation for five-cell pools of lymphocytes (left) and NRBC candidates (right). **B**. An MDS plot for the RNA-seq count data of twelve single lymphocytes (black) and 165 single NRBC candidates (red). Five cells estimated as NRBCs in the primitive stage of their maturation are circled. **C**. A heatmap diagram of 13 marker genes for the three maturation stages of NRBC for lymphocytes and NRBC candidates. The names of five NRBC candidates estimated to be in the primitive stage of their maturation are shown in blue. **D**. The metrics of RNA-seq and WGS libraries prepared by the G&T-seq method from NRBC candidates. The number of genes detected to be expressed by RNA-seq analysis (top), mapping rate to the human reference genome sequence of WGS libraries (middle), and WGS library yield relative to the average yield of WGS libraries from single lymphocytes (bottom) are bar-plotted for five primitive-stage cells (blue) and nine mature-stage cells (black).

In the MDS plot for the gene expression data, five NRBC candidate cells (circled) clustered between the lymphocyte cluster and the cluster of 160 NRBC candidate cells along the dimension-1 axis (**Fig. 3B**). Four out of 12 lymphocyte cells, five primitive NRBC candidates, and nine cells from 160 NRBC cells were visulaized for the expression levels of 13 genes: three primitive-stage, six intermediate-stage, and four mature-stage marker genes of NRBC maturation. [18] Five cells, C1_NRBC_1cell_13, C2_NRBC_1cell_05, C2_NRBC_1cell_25, C4_NRBC_1cell_03, and C4_NRBC_1cell_41, expressed the primitive stage marker *GYPC* and either *CD36* or *CD47* (**Fig. 3C**, **Table S2**). [18] The expression patterns of 13 marker genes observed in the NRBC candidate cells indicate that five cells represent primitive NRBCs, while the other 160 NRBC candidates are enriched with mature-stage cells (**Table S2**).

The number of expressed genes, yield of the WGA-based WGS library, and mapping rate of WGS reads to the reference genome were assessed for NRBC candidate cells. In the five primitive NRBC cells, the number of expressed genes was higher (ranging from 875 to 2,073) than in the other 160 cells (ranging from 24 to 736; average 259; standard deviation 122) (**Fig. 3D**). In the C2_NRBC_05 andC2_NRBC_25 cells, the yields of the WGA-based WGS library and the mapping rates of WGS reads to the reference genome were comparable to those of the lymphocyte-derived WGA-based WGS libraries (**Fig. 3D**, **Table S1**), suggesting the intactness of nuclear genomic DNA in these primitive cells.

## 4. DISCUSSION

Recent studies have shown the feasibility of cell-based NIPT targeting circulating fetal trophoblasts [10-13] and fetal nucleated red blood cells [14] in maternal blood. These studies have also demonstrated the current technical limitations in isolating enough fetal cells with high purity at a low cost and in amplifying genomic DNA without allelic dropout. [9] Improvement in the efficiency of fetal cell isolation is expected to lower the cost of cell-based NIPT for monogenic disorders and facilitate its clinical application.

We previously established a protocol to isolate fetal NRBCs from maternal blood using FACS with erythrocyte-associated surface antigen markers. [15] One advantage of our FACS-based approach is that it does not require specialized devices for NRBC isolation, such as cell scanning systems adopted by other groups. [10, 11] [13, 14] However, the efficiency of the whole-genome amplification of isolated single cells was not consistently sufficient for subsequent whole-genome sequencing. The quality of genomic DNA may have deteriorated because of enucleation and/or apoptosis in most isolated fNRBCs. In our previous study, [15] the mapping rates of single-cell WGS data for NRBC candidates isolated from maternal blood ranged from 20.28% to 90.61%. It has been speculated that NRBCs are present in maternal blood at various maturation stages.

In the present study, we hypothesized that NRBCs in the primitive stage of maturation maintain intact genomic DNA, which is a suitable target for cell-based NIPT. The data obtained for candidate NRBCs from cord blood support our hypothesis that cells in the primitive stages of NRBC maturation maintain intact genomic DNA. In this study, G&T-seq was effective in identifying primitive-stage cells that maintained high-quality DNA among the single NRBC candidates isolated by FACS. Among the 165 candidate NRBCs, we identified five (5/165, 3%) in the primitive stage of maturation based on their gene expression patterns. By conducting WGA-based WGS analysis of the five cells, we confirmed that two NRBCs (2/165, 1.2%) maintained intact genomic DNA suitable for genomic diagnosis. These two cells expressed the three primitive-stage marker genes *GYPC, CD47*, and *CD36*. The expression levels of these three genes will serve as criteria for identifying NRBCs in the primitive stage of maturation using the G & T-seq approach. When we apply our G&T-seq approach to isolate fetal NRBCs from maternal blood, it is expected that it will be possible to distinguish the origins of the isolated NRBC candidates, fetal or maternal, based on the expression levels of hemoglobin F (HBG1 and HBG2) and hemoglobin A (HBB).

As we have shown in the utility of G&T-seq in this study, the application of single-cell multiomics technology is expected to facilitate the development of fNRBC-based NIPT.

## ACKNOWLEDGEMENTS

This work was supported by the National Center for Child Health and Development of Japan (grant numbers 2023B-8 to KN and 2022A-3 to KH) and MEXT KAKENHI (grant number JP23K15853 to NI).

## HUMAN RIGHTS STATEMENT AND INFORMED CONSENT, AND ETHICAL APPROVAL

All procedures were conducted in accordance with the ethical standards of the Institutional Ethical Committee on Human Experimentation (institutional and national) and the Declaration of Helsinki of 1964 and its later amendments. This study was approved by the Institutional Review Board of the National Research Institute for Child Health and Development (IRB number 699). Written informed consent was obtained from all patients.

## DATA AVAILABILITY

Raw sequencing data (WGS and RNA-seq) are available upon request from the corresponding author (KN).

## LIST OF THE SUPPORTING INFORMATION

Table S1: The metrics of G&T-seq libraries from lymphocytes and NRBCs

Table S2: Transcript per million (TPM) values of 13 markers genes in single lymphocytes and NRBC candidates

